# Health Literacy and Lifestyle Scores Among a Small but Diverse Group of Older Asians Who Attended Community Health Events in Los Angeles

**DOI:** 10.64898/2026.05.27.26354181

**Authors:** Enxi Zhang, Tricia Tran, Kaela Shun, Darcey Tran, Abigail Tsai, Emma Kwang, Maral DerSarkissian, Tony Kuo

**Affiliations:** University of California, Los Angeles (UCLA); Institute for Society and Genetics, UCLA; Department of Ecology and Evolutionary Biology, UCLA; Department of Integrative Biology and Physiology, UCLA; Department of Psychology, UCLA; Analysis Group, Inc.; Department of Epidemiology, UCLA Fielding School of Public Health

## Abstract

The Asian population in Los Angeles is among the largest and most heterogeneous in the U.S. This is true culturally and health-wise. Older Asians have differing risks for cardiovascular and cardiometabolic disease, depending on their ethnicity, health literacy, and lifestyle choices. This pilot examines several of these factors in a small but diverse group of older Asian adults who attended community health events from 2024-2025. Self-reported and biometric data were collected at five such events hosted by the Asian Pacific Health Corps at UCLA. The pilot generated health literacy and lifestyle (HLL) scores for all participating attendees and explored how they relate to their socio-demographics, healthcare habits, and predictions of their own health data. Overall, there were significantly more females than males with higher HLL scores (p = 0.027). College education (p = 0.028) and “normal” ranges for biometric data (e.g., blood pressure, BMI, blood glucose, cholesterol) were related to higher median HLL scores. With a few exceptions, fewer than 50% accurately predicted their biometric numbers regardless of HLL scores, suggesting a disconnect between perception and reality, and that better provider-patient communication may help foster greater patient understanding about their chronic conditions. These HLL score distributions indicate that educational attainment, better awareness of one’s health, and high health literacy are individual factors that may influence older Asians’ understanding and potential approach to managing their health conditions.

## Introduction

Older Asians are the fastest growing population in the U.S. Yet, they are underrepresented in health-related research. This may be due to a variety of factors such as a lack of participation in clinical trials, language barriers, and immigration challenges.^1^ Disparities by ethnicity (Chinese/Korean/Vietnamese, etc.) and other characteristics unique to these subgroups are often understudied.^2^ Even pilot projects about them are sparse in the literature. For example, gaps in health literacy and lifestyle choices—cardiovascular and cardiometabolic risk factors that are often amenable to behavioral modification (e.g., through physical activity promotion, better sleep hygiene, nutritious diet)^3^—are particularly prevalent.

We address these gaps in health practices by conducting a pilot analysis of a small but diverse group of older Asian ethnic adults who attended five community health events in Los Angeles. The goal of the analysis was to gain a better understanding of how and if high health literacy may lead to better knowledge and predictions of one’s own health. In this pilot, health literacy is defined as the ability to understand and use health information to make decisions on one’s health.^4^

## Methods

We collected survey and biometric data—blood pressure (mm Hg via electronic cuffs, American Heart Association [AHA] guidelines), body mass index (BMI, using Asian cut-points), body fat percentage (based on age and sex), blood glucose (mg/dL), and cholesterol (mg/dL)—from 56 older Asian adults (age >50+) who attended five local community health events during October 2024 to May 2025. These Los Angeles events were hosted by the student-run Asian Pacific Health Corps at the University of California, Los Angeles (UCLA), with support from local community health centers. All 56 attendees who participated in the pilot received health screenings and other services at the event. They consented to participating anonymously and completed a survey and required biometric measurements. The survey asked about socio-demographic information (age, sex, ethnicity, income, education), personal and family health history, routine healthcare habits, ability to obtain healthy food, and whether they have health insurance. A separate set of questions captured information about health literacy and lifestyle (HLL) choices based on AHA criteria; this information was then used to generate a numeric score for each attendee. Lifestyle questions included: smoking (daily use), secondhand smoke exposure (household member who smokes), weekly hours of moderate intensity exercise, weekly alcohol consumption, bedtime and nightly hours of sleep, and daily fruit and vegetable consumption. All tools (survey, HLL questions) were available in multiple languages, including English, Korean, simplified and traditional Chinese, Spanish, and Vietnamese. Interpreters were also present at these events to assist. The survey took approximately 10 minutes to complete and biometric data were linked to self-reported information via a randomly assigned number.

HLL scores were tabulated based on how well each participating attendee responded to the AHA’s scoresheet—1 point each for knowing and fulfilling the recommended behaviors, 0.5 points for coming close, and 0 points for not knowing. Above and below median thresholds were calculated and dichotomized to compare these scores by socio-demographics and other variables, including self-prediction of health profile. We employed two-tailed Z tests to assess statistical significance (threshold: p < 0.05).

## Results

The median HLL score for the 56 participating attendees was 8 out of 12 points possible; a score ≥8 was considered “high health literacy,” while a score <8 was considered “low health literacy.” **Table 1** reports on socio-demographics and health behaviors stratified by above versus below median HLL score. Of the 56 attendees, 42.9% identified as Chinese, 3.6% Filipino, 3.6% Japanese, 8.9% Korean, 12.5% Taiwanese, and 14.3% Vietnamese. There were no statistically significant differences in median HLL scores among these various Asian subgroups. While the median HLL scores by socio-demographics and healthcare habits showed differences, these observations did not reflect the attendees’ self-predictions of their own biometric health data (their predictions were generally inaccurate; see **Table 1, Table 2**, and **Figure 1**). Those with high median HLL scores, for example, tended to be female (70.6%, p = 0.027), college educated (47.1%, p = 0.028), and visited their doctor routinely (73.0%, albeit p = 0.139). With a few exceptions, fewer than 50% accurately predicted their biometric numbers regardless of HLL scores.

**Table 1.** Comparison of above versus below median Health Literacy and Lifestyle (HLL) score among older Asian attendees of community health events in Los Angeles by.

| **n=56** | **Above Median**<br>**HLL Score ($\geq 8$)**<br>**(n=34)** | **Below Median**<br>**HLL Score**<br>**($< 8$) (n=22)** | **p-value** |
| --- | --- | --- | --- |
| **Age (years)** | Mean (standard deviation) | Mean (standard deviation) |  |
|  | 60.1 (18.63) | 55.7 (19.40) |  |
|  | n (%) | n (%) |  |
| **Sex** |  |  |  |
| Female | 24 (70.6) | 9 (40.9) | 0.027 |
| Male | 8 (23.5) | 7 (31.8) | 0.494 |
| No response | 2 (5.9) | 6 (27.3) | 0.025 |
| **Race/ethnicity** |  |  |  |
| Chinese | 14 (41.2) | 10 (45.5) | 0.752 |
| Filipino | 2 (5.9) | 0 | 0.247 |
| Japanese | 2 (5.9) | 0 | 0.247 |
| Korean | 3 (8.8) | 2 (9.1) | 0.973 |
| Taiwanese | 4 (11.8) | 3 (13.6) | 0.836 |
| Vietnamese | 5 (14.7) | 3 (13.6) | 0.911 |
| Other | 1 (2.9) | 0 | 0.417 |
| Decline to state | 0 (0) | 1 (4.5) | 0.210 |
| No response | 3 (8.8) | 3 (13.6) | 0.570 |
| **Income** |  |  |  |
| None | 3 (8.8) | 4 (18.2) | 0.301 |
| < \$20,000 | 1 (2.9) | 2 (9.1) | 0.301 |
| \$20,000 - \$39,999 | 6 (17.6) | 2 (9.1) | 0.372 |
| \$40,000 - \$89,999 | 7 (20.6) | 3 (13.6) | 0.507 |
| > \$90,000 | 2 (5.9) | 2 (9.1) | 0.828 |
| Decline to state | 15 (44.1) | 9 (40.9) | 0.813 |
| **Education** |  |  |  |
| Below high school | 0 (0) | 2 (9.1) | 0.073 |
| High school | 4 (11.8) | 4 (18.2) | 0.503 |
| College | 16 (47.1) | 4 (18.2) | 0.028 |
| Above college | 6 (17.6) | 5 (22.7) | 0.640 |
| Technical/vocational school | 0 (0) | 1 (4.5) | 0.210 |
| Decline to state | 8 (23.5) | 6 (27.3) | 0.363 |
| **Personal and/or family health history** |  |  |  |
| Cardiovascular disease | 5 (14.7) | 8 (36.4) | 0.248 |
| Other chronic conditions (e.g., obesity, hypertension, diabetes) | 20 (58.8) | 7 (31.8) | 0.048 |
| Not applicable | 9 (26.5) | 7 (31.8) | 0.665 |
| **Visit doctor routinely** |  |  |  |
| Yes | 30 (88.2) | 16 (72.7) | 0.139 |
| No | 3 (8.8) | 6 (27.3) | 0.066 |
| No response | 1 (2.9) | 0 (0) | 0.417 |
| **Health insurance** |  |  |  |
| Yes | 24 (70.6) | 16 (72.7) | 0.863 |
| No | 6 (17.6) | 3 (13.6) | 0.690 |
| No response | 4 (11.8) | 3 (13.6) | 0.836 |
| **Able to communicate health concerns to doctor?** |  |  |  |
| Yes | 22 (64.7) | 10 (45.5) | 0.155 |
| No | 8 (23.5) | 7 (31.8) | 0.494 |
| No response | 4 (11.8) | 5 (22.7) | 0.275 |
| **Struggle for healthy food or need government assistance?** |  |  |  |
| Currently | 7 (20.6) | 5 (22.7) | 0.849 |
| In the past | 1 (2.9) | 1 (4.5) | 0.752 |
| No | 25 (73.5) | 13 (59.1) | 0.259 |
| No response | 1 (2.9) | 3 (13.6) | 0.129 |

**Table 2.**
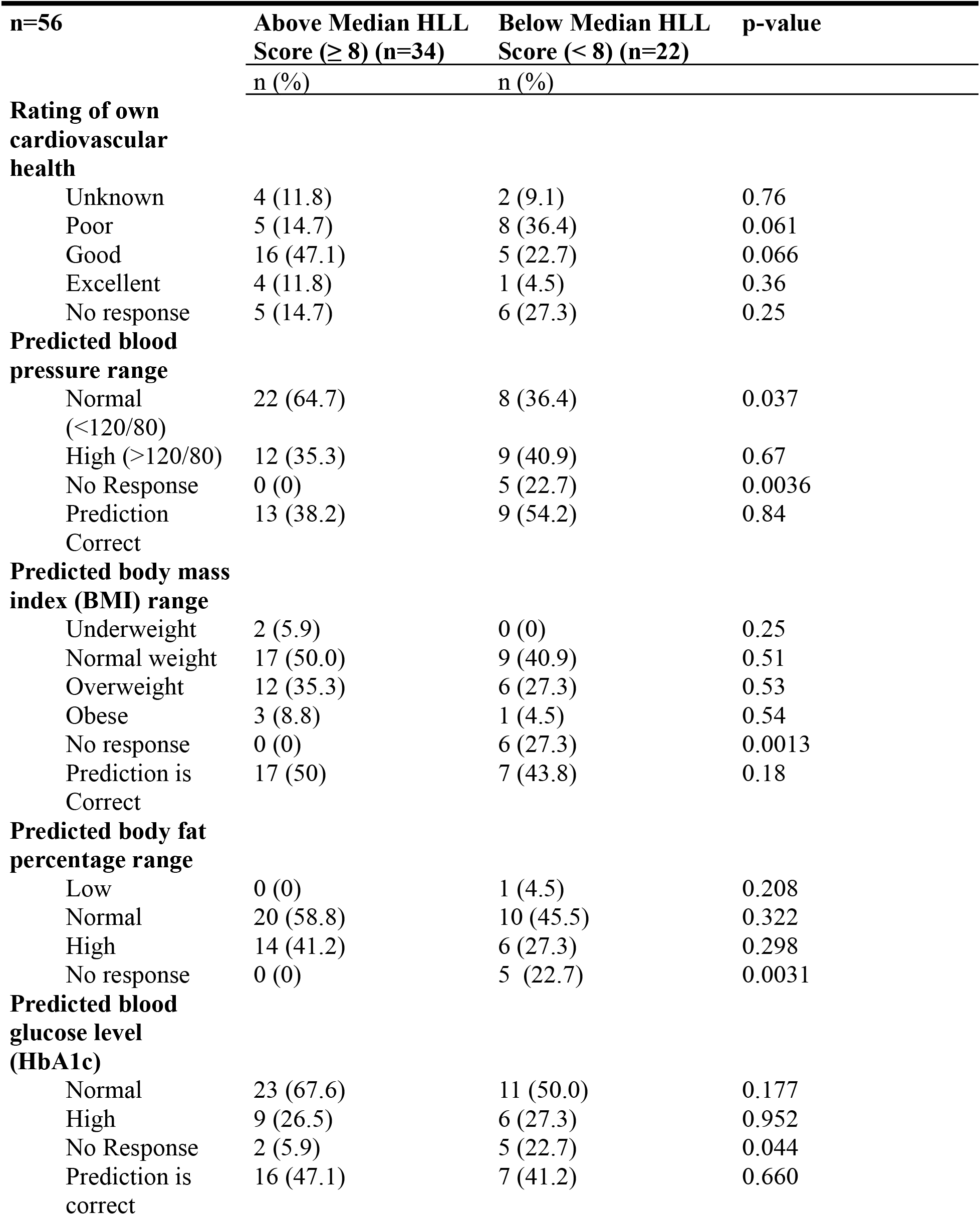
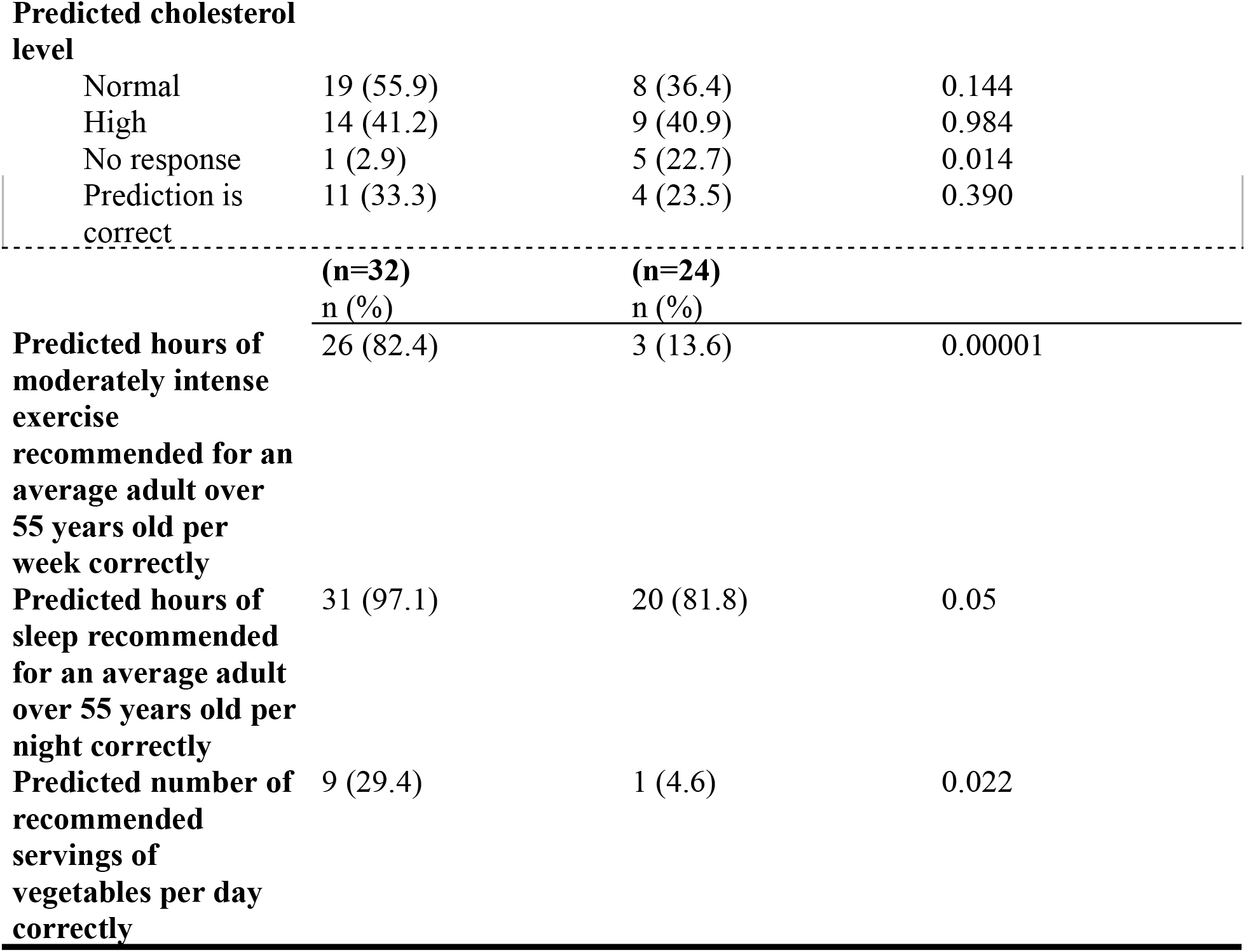
Comparison of above versus below median Health Literacy and Lifestyle (HLL) score among older Asian attendees of community health events in Los Angeles by self-ratings and self-predicted indicators on health and health behaviors.

| **n=56** | **Above Median HLL**<br>**Score ($\geq 8$) (n=34)** | **Below Median HLL**<br>**Score ($< 8$) (n=22)** | **p-value** |
| --- | --- | --- | --- |
|  | **n (%)** | **n (%)** |  |
| **Rating of own cardiovascular health** |  |  |  |
| Unknown | 4 (11.8) | 2 (9.1) | 0.76 |
| Poor | 5 (14.7) | 8 (36.4) | 0.061 |
| Good | 16 (47.1) | 5 (22.7) | 0.066 |
| Excellent | 4 (11.8) | 1 (4.5) | 0.36 |
| No response | 5 (14.7) | 6 (27.3) | 0.25 |
| **Predicted blood pressure range** |  |  |  |
| Normal (<120/80) | 22 (64.7) | 8 (36.4) | 0.037 |
| High (>120/80) | 12 (35.3) | 9 (40.9) | 0.67 |
| No Response | 0 (0) | 5 (22.7) | 0.0036 |
| Prediction Correct | 13 (38.2) | 9 (54.2) | 0.84 |
| **Predicted body mass index (BMI) range** |  |  |  |
| Underweight | 2 (5.9) | 0 (0) | 0.25 |
| Normal weight | 17 (50.0) | 9 (40.9) | 0.51 |
| Overweight | 12 (35.3) | 6 (27.3) | 0.53 |
| Obese | 3 (8.8) | 1 (4.5) | 0.54 |
| No response | 0 (0) | 6 (27.3) | 0.0013 |
| Prediction is Correct | 17 (50) | 7 (43.8) | 0.18 |
| **Predicted body fat percentage range** |  |  |  |
| Low | 0 (0) | 1 (4.5) | 0.208 |
| Normal | 20 (58.8) | 10 (45.5) | 0.322 |
| High | 14 (41.2) | 6 (27.3) | 0.298 |
| No response | 0 (0) | 5 (22.7) | 0.0031 |
| **Predicted blood glucose level (HbA1c)** |  |  |  |
| Normal | 23 (67.6) | 11 (50.0) | 0.177 |
| High | 9 (26.5) | 6 (27.3) | 0.952 |
| No Response | 2 (5.9) | 5 (22.7) | 0.044 |
| Prediction is correct | 16 (47.1) | 7 (41.2) | 0.660 |
| **Predicted cholesterol level** |  |  |  |
| Normal | 19 (55.9) | 8 (36.4) | 0.144 |
| High | 14 (41.2) | 9 (40.9) | 0.984 |
| No response | 1 (2.9) | 5 (22.7) | 0.014 |
| Prediction is correct | 11 (33.3) | 4 (23.5) | 0.390 |

|  | **(n=32)**<br>n (%) | **(n=24)**<br>n (%) |  |
| --- | --- | --- | --- |
| **Predicted hours of moderately intense exercise recommended for an average adult over 55 years old per week correctly** | 26 (82.4) | 3 (13.6) | 0.00001 |
| **Predicted hours of sleep recommended for an average adult over 55 years old per night correctly** | 31 (97.1) | 20 (81.8) | 0.05 |
| **Predicted number of recommended servings of vegetables per day correctly** | 9 (29.4) | 1 (4.6) | 0.022 |

## Discussion

This pilot analysis of a small but unique group of older Asian adults, representing unique ethnicities, contributes valuable information and insights into how health screenings and related services could be improved for many of these underserved individuals. Increasing health literacy, raising awareness of personal health, and facilitating healthy lifestyle practices can all lead to a better understanding of health conditions and ways in which these older adults could reduce their risks for cardiovascular and cardiometabolic diseases. The pilot’s results also speak to a potential need for communicating health concerns in a culturally relevant way and in recognizing the slight differences in physiology (e.g., BMI or body fat percentage cut-points) among these Asian ethnicities.^5,6^ The above versus below median HLL scores offer interesting delineations on which of these two groups may be ready for coaching or tailored provider-patient communications that can accelerate their health knowledge and nudge them toward more self-empowered management of their chronic conditions.

### Limitations

The most notable limitations of this project include the following. (i) It is a pilot analysis, with a small convenience sample of 56 recruited from five community health events in Los Angeles, which limits generalizability. (ii) The survey and biometric data, while insightful in their presentation, given the diverse ethnicities they represent (the pilot’s strength), nevertheless lacked the power to make causal comparisons/inferences by subgroups. Lastly, (iii), self-reported data, unlike biometric data, were likely affected by selection and social desirability biases.

## Data Availability

All data produced in the present study are available upon reasonable request to the authors

## Future Steps

The Asian Pacific Health Corps plans to use these pilot data to improve their year-round health event planning and programming for older Asians in Los Angeles. They are presently applying the data to help develop practical strategies for promoting health literacy and self-care among these unique ethnic groups.

