## Supplementary material for "Health Literacy and Lifestyle Scores Among a Small but Diverse Group of Older Asians Who Attended Community Health Events in Los Angeles": Figure 1

**Figure 1: Boxplots of Health Literacy and Lifestyle (HLL) scores for four health metrics among older Asian attendees of community health events in Los Angeles – blood pressure, body mass index (BMI), blood glucose level, and cholesterol**

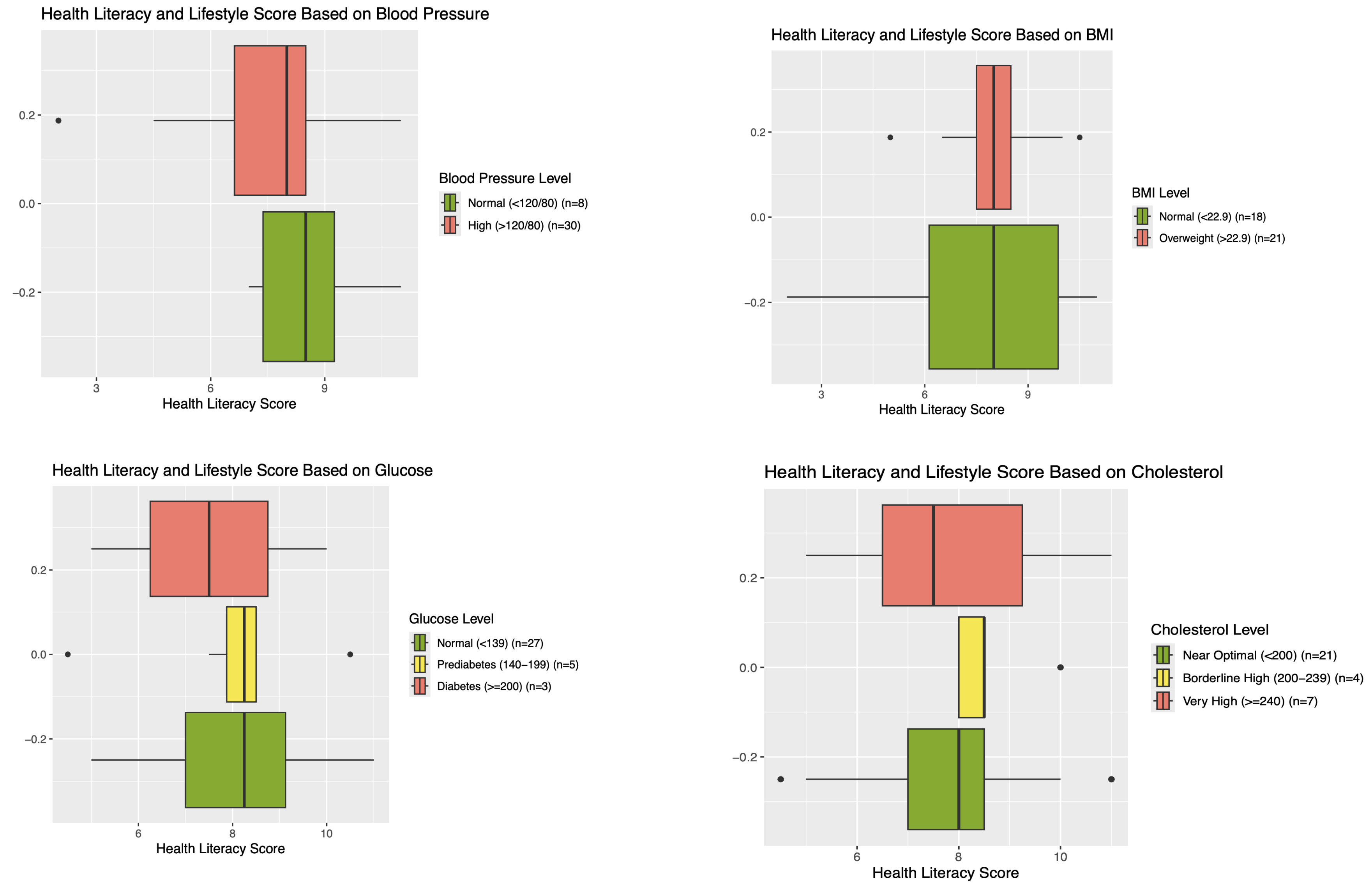
